# The Effects of Virtual Reality Neuroscience-based Therapy on Clinical and Neuroimaging Outcomes in Patients with Chronic Back Pain: A Randomized Clinical Trial

**DOI:** 10.1101/2023.07.24.23293109

**Authors:** Marta Čeko, Tassilo Baeuerle, Lynn Webster, Tor D. Wager, Mark A. Lumley

## Abstract

Chronic pain remains poorly managed. The integration of innovative immersive technologies (i.e., virtual reality (VR)) with recent neuroscience-based principles that position the brain as the key organ of chronic pain may provide a more effective pain treatment than traditional behavioral therapies. By targeting cognitive and affective processes that maintain pain and potentially directly changing neurobiological circuits associated with pain chronification and amplification, VR-based pain treatment has the potential for significant and long-lasting pain relief.

We tested the effectiveness of a novel VR neuroscience-based therapy (VRNT) to improve pain-related outcomes in n = 31 participants with chronic back pain, evaluated against usual care (n = 30) in a 2-arm randomized clinical trial (<u>NCT04468074)</u>. We also conducted pre- and post-treatment MRI to test whether VRNT affects brain networks previously linked to chronic pain and treatment effects. Compared to the control condition, VRNT led to significantly reduced pain intensity (g = 0.63) and pain interference (g = 0.84) at post-treatment vs. pre-treatment, with effects persisting at 2-week follow-up. The improvements were partially mediated by reduced kinesiophobia and pain catastrophizing. Several secondary clinical outcomes were also improved, including disability, quality of life, sleep, and fatigue. In addition, VRNT was associated with modest increases in functional connectivity of the somatomotor and default mode networks and decreased white matter fractional anisotropy in the corpus callosum adjacent to anterior cingula, relative to the control condition. This, VRNT showed preliminary efficacy in significantly reducing pain and improving overall functioning, possibly via changes in somatosensory and prefrontal brain networks.

## Introduction

Chronic musculoskeletal pain remains poorly managed, with leading behavioral interventions having generally small benefits [28,31,63,92]. The limited efficacy of behavioral approaches may stem from the fact that they are not guided by a conceptual model that places the brain as the centerpiece of the chronic pain experience, do not emphasize the possibility of reducing pain by changing cognitive and affective processes that drive pain, and may lack powerful experiential exercises to change those thoughts and feelings [51].

The integration of innovative immersive technologies with recent neuroscience-based behavioral approaches may provide a more effective pain treatment with potentially broader reach [4,35,52,86,94]. For example, Virtual reality (VR) can reduce pain during medical procedures and hospitalization, presumably by providing immersive distraction from pain [29,35,70,73]. More recently, VR-based interventions have shown preliminary efficacy as adjunctive [62] or stand-alone treatments for chronic or persistent pain [21,22,55]. Crucially, and in contrast to acute pain reduction afforded by distraction-based VR implementations, VR used for chronic pain has the potential for long-lasting pain relief [30,86], likely reducing fearful beliefs about the bodily danger of pain and enhancing attributions of brain-based control of pain. Such cognitive and affective changes might be reflected in changes in neurobiological circuits associated with pain chronification and amplification [5,9,61,63].

We created a multimodal, self-management program for chronic back pain (CBP), ‘Virtual Reality Neuroscience-based Therapy’ (VRNT) to test the effect on clinical pain and brain outcomes. VRNT integrates contemporary pain and affect neuroscience focusing on the brain as the key determinant of chronic pain experience through predictive processing by integrating nociceptive input with cognitive, emotional, and social factors [5,23,43,61]. In addition to a pain neuroscience education component, VRNT includes cognitive, behavioral, and affective exercises and experiences in the immersive virtual environment to increase real-time awareness of how the brain processes pain and decrease learned maladaptive cognitions and emotions, including catastrophizing and fear-related avoidance. Thus, VRNT goes beyond traditional therapies to address central drivers of chronic pain in an immersive, experiential format, targeting meaningful and durable pain reduction.

Chronic musculoskeletal pain has been associated with brain changes that reflect sensitization and neuroplasticity in distributed neural circuits [2,6,8–10,15,26,38,39,42,43,45,49,50,57,58,71,80,87] involved in somatomotor processes and sensory discrimination (‘somatomotor network’), self-reference, motivation and emotion appraisal (‘default mode network’, DMN; fronto-striatal circuitry), and attention allocation to salient stimuli (‘cingulo-opercular network’). Limited evidence suggests that these brain changes could be partially reversed with successful behavioral treatments [4,33,44,68,81,82], supporting the notion that pain-related maladaptive plasticity within these distributed large-scale neural networks could be normalized if pain is improved [65].

The primary objective of this study was to assess the effectiveness of the VRNT program to reduce pain and improve functioning in patients with CBP. We hypothesized that VRNT would reduce pain intensity and interference compared to the control waitlist condition, with reductions in pain catastrophizing and kinesiophobia mediating the effects. We also conducted pre- and post-treatment MRI and hypothesized that VRNT would affect structural and functional connectivity in the abovementioned brain networks associated with chronic pain and treatment effects.

## Methods

### Participants

Adults with CBP were recruited and enrolled via on-line advertising (Facebook) from the Boulder/Denver Colorado Metro Area. Inclusion criteria were aged 21 to 70, experiencing back pain on at least half the days of the last 6 months, and the average pain intensity over the last week of at least 4 out of 10 at study entry. We excluded people with leg pain worse than back pain (to exclude those with neuropathic pain); other chronic pain besides CBP; major medical and neurological disorders; mental health disorders not controlled with medication; a history of substance abuse; pain-related compensation or litigation in the past year; a history of vertigo, dizziness, susceptibility to motion sickness, head injury within 6 months, (digital) eye strain, or computer vision syndrome; and standard MRI contraindications as determined by an MRI safety screen.

The institutional review board of the University of Colorado Boulder approved the study, and all participants provided written consent (via Docusign). The study was pre-registered on ClinicalTrials.gov (Identifier: <u>NCT04468074</u>). Study enrollment began in June 2020; randomization began in July 2020; and last follow-up was completed in June 2021. The study followed the Consolidated Standards of Reporting Trials (CONSORT) guidelines for social and psychological intervention, (Fig 1.; [56]).

**Figure 1.**
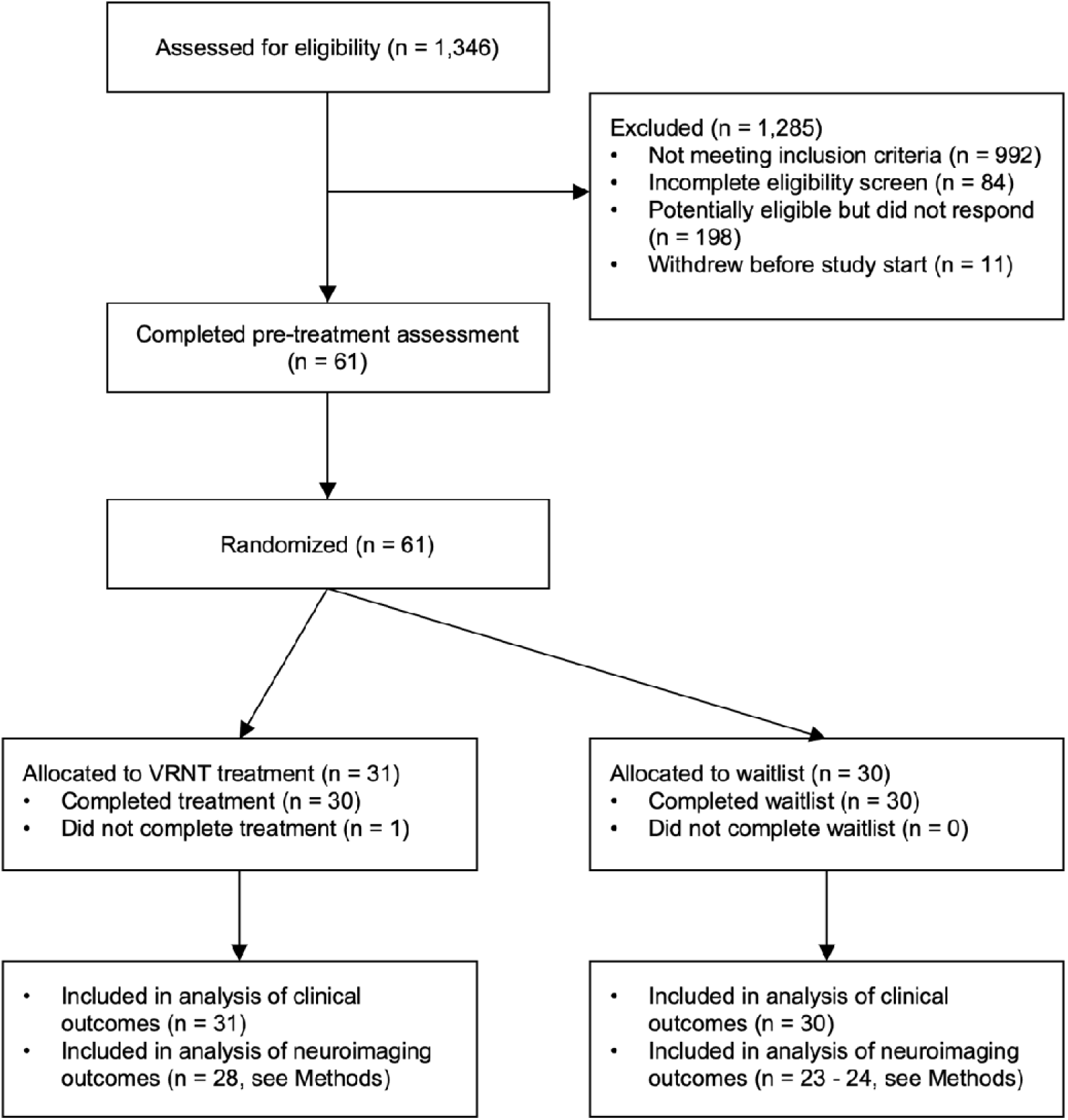
CONSORT Participant flow diagram.

### Procedure

The study consisted of an 8-week VRNT program (or waitlist/usual care control), with outcome measures (self-report measures, MRI) assessed at multiple timepoints. All self-report measures were completed at home on-line (via Qualtrics), including at pre-treatment, mid-treatment, post-treatment, and follow-up. Pre-treatment assessment was completed by all participants prior to randomization and consisted of two administrations of most primary and secondary self-report measures: at study entry (week -2) and again at week 0; the latter also included an MRI session (Fig. 2A). After the MRI, participants were randomized by a research assistant (using Python 3.7) to one of the two conditions (VRNT or Control) in a 1:1 ratio, stratified by sex and age; all participants continued to receive their usual care. Participants assigned to VRNT began the intervention immediately, whereas those assigned to the Control condition were offered VRNT after the follow-up assessment. All participants were assessed for self-reported outcomes at week 4 (mid-way through VRNT/Control), at post-treatment (week 8), and at 2-week follow-up (week 10). The MRI was repeated at post-treatment (week 8). Research staff administering the MRI and analyzing data were blinded to condition assignment; participants were instructed not to discuss their assignment during these sessions. Research staff involved in recruitment, enrollment and randomization were not involved in data analysis.

**Figure 2.**
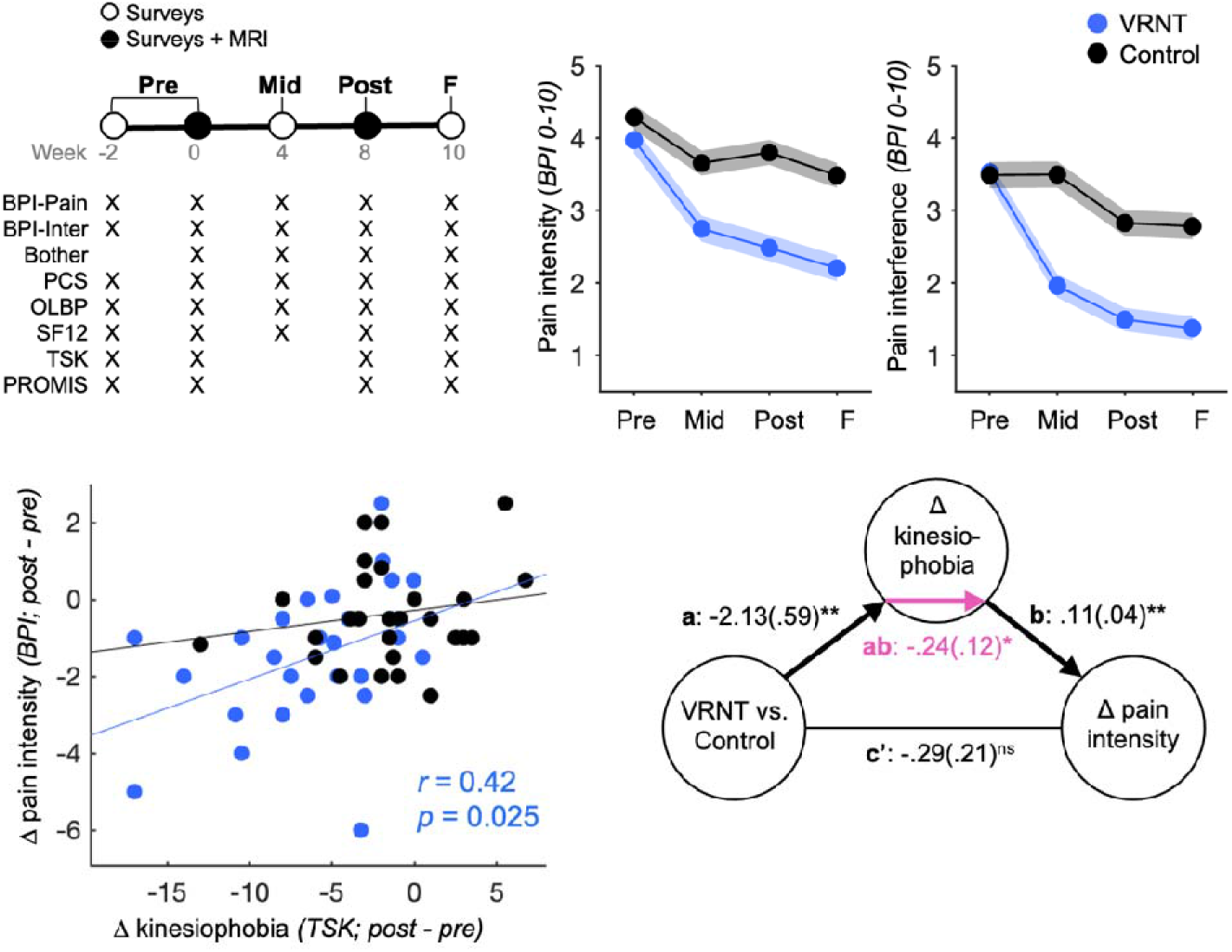
Treatment effects on behavioral outcomes. **(A)** The study design consisted of 5 assessments, including on-line questionnaires administered at week -2 and 0 (averaged as pre-treatment timepoint scores), at week 4 (mid-treatment timepoint), at week 8 (post-treatment timepoint) and at week 10 (follow-up timepoint), and two MRI sessions (at weeks 0 and 8). **(B)** Plot shows pain intensity levels per timepoint; shading indicates standard error. **(C)** Plot shows pain interference levels per timepoint; shading indicates standard error. **(D)** Scatter plot shows the relationship between the pain intensity post-pre difference score and the kinesiophobia post- pre difference score for each individual. **(E)** Mediation analysis testing whether the kinesiophobia post-pre difference score mediates the relationship between condition assignment (VRNT, Control) and the pain intensity post-pre difference score. Mediation coefficients tested for significance using 10,000 bootstrap samples. * p < .05; ** p < .01; *** p < .0001; two-sided tests; BPI, Brief Pain Inventory, BPI-Pain, pain intensity; BPI-Inter, pain interference; Bother, bothersomeness; PCS, Pain Catastrophizing Scale; OLBP, Oswestry Low Back Pain Questionnaire; SF12, Short-Form Quality of Life Questionnaire; TSK, Tampa Scale of Kinesiophobia; PROMIS, Patient Reported Outcome Measurement Information System; Pre, pre-treatment; Mid, mid-treatment; Post, post-treatment; F, follow-up; VRNT, Virtual Reality Neuroscience-based Therapy.

### Intervention: Virtual Reality Neuroscience-based Therapy (VRNT)

The VRNT program was initiated by a 34-minute educational video (presented via Zoom), which introduced the neuroscience of acute and chronic pain, drivers of chronic pain (e.g., catastrophizing, stress, fear / fear-avoidance cycle), pain triggers, and the scientific principles behind VRNT. The study coordinator then worked with each VRNT participant to personalize the VRNT application by selecting a 3-dimensional avatar (from 9 different avatar options of male/female/neutral gender and body shapes/sizes), and creating a 3D-animated, audiovisual representation of the participant’s own pain experiences (shape, color, sound, animation) in the translucent avatar see Supplementary Fig.1 for an example). This final customized representation was loaded into a mobile VR device and provided to the participant to use at home for the 8 weeks. The Mobile VR device was a Samsung GearVR (2017), which included a head-mounted display, a hand controller and a Samsung Galaxy S-9 phone. Participants were asked to complete 2 VRNT modules per day, 5 days per week, for a total of 40 days of module completion. A calendar prescribed the sequence of modules to be completed but allowed the participant to choose some of the sessions. An accompanying workbook provided background on each module. In addition, weekly calls served to answer participants’ questions, obtain feedback on VRNT modules, and assess and encourage program adherence.

The content of the VRNT program was organized into 17 different exercises or “modules” of varying duration (daily time for 2 modules: 7 to 27 minutes; average 20 minutes). Each module had a pre-recorded voiceover, providing education and/or guiding the participant through the exercise while wearing the headset. Overall, the modules covered 8 categories, each with various techniques or exercises: 1) Pain education (pain neuroscience and skills); 2) Relaxation and mindfulness training (mindful scanning of the body and mindful detachment from thoughts and sensations); 3) Interoception (identifying and labeling internal, usually somatic experiences); 4) Mindful escapes, passive distraction, and shifting focus (distracting from pain and shifting focus toward or away from pain to improve attention control in real-world situations); 5) Thought appraisal (tracking and evaluating automatic thoughts about pain); 6) Diaphragmatic breathing (immersive breath training to support self-regulation and relaxation); 7) Graded exposure therapy (hierarchical exposure of patients to their own pain, to pain triggers such as cold temperature, and to animated movements that commonly cause pain); and 8) Emotion triggers (improving awareness and self-regulation of emotions). Although VRNT incorporates some techniques from common behavioral pain management approaches (e.g., Cognitive Behavioral Therapy (CBT)), it also utilizes techniques from newer therapies, such as Pain Reprocessing Therapy (PRT) and Emotional Awareness and Expression Therapy (EAET), which have been shown to yield substantial pain reductions in several trials [4,52,94]. Consistent with these newer models, VRNT explicitly teaches participants about the reversibility of the central neuroplastic changes associated with pain chronification. In contrast, most chronic pain behavioral therapies view pain as chronic of unknown origin and less changeable [51]. VRNT also augments classic cognitive (neuroscience, skills) education with experiential learning by guiding the participant through self-discovery of how their own brain processes pain (e.g., sensations, thoughts, emotions); to this VRNT leverages interoceptive modeling techniques in which the participant’s own animated VR pain representation changes during the modules.

### Measures

Following the recommendations of the Initiative on Methods, Measurement, and Pain Assessment in Clinical Trials [17] and National Institutes of Health Pain Consortium’s Report on Research Standards for Chronic Low Back Pain [16], we assessed multiple domains of self-report outcome measures. We had two primary outcomes (pain intensity and interference) and numerous secondary outcomes. All the outcomes and potential mediators listed below (except pain bothersomeness) were assessed twice at pretreatment (weeks 0 and 2), and we averaged the two assessments to make a more reliable pre-treatment value of each measure.

### Primary outcomes

#### Pain intensity

We used the single item (“average pain intensity over the past week”), rated 0 to 10, from the Short-Form Brief Pain Inventory (BPI-SF [14]. In addition to analyzing the condition means on this item, we also determined a *clinically meaningful improvement* as a reduction in pain intensity of at least 30% from pre-treatment (post-treatment minus pre-treatment) and *substantial pain reduction* as at least 50% from pre-treatment [84,95].

#### Pain interference

We averaged the ratings (0 to 10) of the 7 items from the BPI-SF pain interference subscale to assess the degree to which participants’ pain interfered with daily life activities over the past week. Again, we analyzed both the condition means and the frequency of clinically meaningful and substantial reductions in interference pre-treatment.

### Secondary outcomes

#### Pain bothersomeness

Back pain bothersomeness over the past week was assessed using a 0-10 rating, as used previously [12].

#### Pain-related disability

The 10 items of the Oswestry Low Back Pain Disability Questionnaire (OLBPQ) [19] were summed.

#### Psychological distress

Short-form measures from the Patient Reported Outcomes Measurement Information System (PROMIS; [47,74] assessed depressive symptoms (form 8a), anxiety (form 8a), and anger (form 5a) over the past week; sums of item ratings were analyzed.

#### Sleep

The PROMIS measure (form 8a) assessed sleep quality over the past week; the sum of items was analyzed.

#### Fatigue

The PROMIS measure (form 8a) assessed fatigue over the past week; the sum of items was analyzed.

#### Quality of life

The Short-Form Health Survey (SF-12 [90] assessed the impact of health on everyday life. The final score was averaged across two subscale scores (mental health, physical health).

### Potential treatment mediators

Two variables were examined as potential mediators of treatment effects on the primary outcomes.

#### Kinesiophobia

The 11-item Tampa Scale of Kinesiophobia (TSK; [83]) assessed fear of movement and beliefs that pain indicates injury. Item ratings were summed.

#### Pain catastrophizing

The 13-item Pain Catastrophizing Scale [75] assessed rumination, magnification, and helplessness related to pain. Item ratings were summed.

Other potential mediators included fear of pain, pain attitudes, self-efficacy, optimism, meaning and purpose in life, mindfulness, and emotion regulation capacity. Results of analyses of these variables are reported in Supplementary Table 1.

### Neuroimaging outcomes

We collected several neuroimaging outcomes to establish brain mechanisms associated with treatment response. Neuroimaging outcomes comprised two types of images: resting-state functional fMRI suitable for analysis of functional connectivity between brain regions, and diffusion-weighted images (DWI) suitable for the analysis of white matter fractional anisotropy (FA), indicative of structural white matter integrity between brain regions.

### MRI acquisition

Neuroimaging outcomes were collected at pre-treatment session 2 and at post-treatment. Whole-brain fMRI data were acquired on a 3T Siemens MAGNETOM Prisma MRI scanner at the Intermountain Neuroimaging Consortium facility at the University of Colorado, Boulder. Structural images were acquired using high-resolution T1 spoiled gradient recall images (SPGR) and were used for anatomical localization and warping to standard MNI space only. Functional images were acquired with a multiband echo-planar imaging (EPI) sequence (TR = 460 ms, TE = 27.2 ms, field of view = 220 mm, multiband acceleration factor = 8, flip angle = 44°, 64 x 64 matrix, 2.7 x 2.7 x 2.7 mm voxels, 56 interleaved ascending slices, phase encoding posterior >> anterior). One resting-state functional run of 6 mins duration was acquired. DWI images were acquired with a single-shot, multiband, multi-shell protocol (TR = 4,000 ms, TE = 77.0 ms, multiband acceleration factor = 3, field of view = 224 mm, 52 x 52 matrix, 2.0 x 2.0 x 2.0 mm voxels, interleaved 72 slices, total duration ∼ 12 mins) with 4 diffusion-weighted shells at b = 2400 s/mm^2^ with varying diffusion directions (44, 47, 42, 40).

### MRI data preprocessing

MRI data were preprocessed using standardized pipelines, with steps summarized in Supplementary Methods. Anatomical and resting-state data were preprocessed using *fmriprep* 21.0.0 [18], which is based on *nipype* 1.6.1 [24]. DWI (white matter) data were preprocessed using QSIPrep [13] and fractional anisotropy (FA) maps were derived using DTIFIT in FSL by fitting a diffusion tensor model in each voxel of the preprocessed DWI data and calculating voxel-wise FA values.

### Statistical analyses

### Sample size estimation

Power analysis targeted 80% power (α = .05) to detect a large effect (f = 0.40) on pain intensity at post-treatment with the pre-treatment intensity as a covariate. A total sample of 52 was needed, and we overrecruited slightly, expecting some attrition.

### Treatment effects on primary and secondary outcomes

Intent-to-treat analyses (including all randomized patients) were performed for primary and secondary outcomes using a mixed-effects model (fitlme, MATLAB 2020b), including a condition by time interaction (VRNT vs. Control x post- vs. pre-treatment), covariates for age and sex, and a random intercept per participant. For further analyses, change scores were post-treatment minus pre-treatment for participants with available pre- and post-treatment data (N = 30 per condition). Treatment effect sizes were calculated as the VRNT vs. Control difference in change scores divided by the pooled standard deviation of change scores, with the Hedge’s *g* correction and bootstrapped (n = 10,000) confidence intervals (mes toolbox [27], MATLAB 2020b). Conditions were compared on frequency of at least 30% and 50% improvement from pre- to post-treatment) on the two primary outcomes via chi-square tests (https://www.socscistatistics.com/tests/chisquare/).

### Potential mediators of treatment

To examine potential mechanisms of VRNT treatment, we first compared the VRNT and Control conditions on change in potential mediators using the same approach as for outcomes (condition by time interaction analyses, controlled for age and sex). Next, we computed Pearson correlations between mediator change and primary outcome change scores for the entire sample, and significant correlations were examined further with a formal mediation analysis, which tested whether a change in the mediator accounted for treatment condition effects on change in each primary outcome (i.e., VRNT vs. Control → Δ potential mediator → Δ primary treatment outcome). Mediation analyses were computed using the Canlab Mediation Toolbox (MATLAB 2020b)[89] Statistical significance of mediation was derived with 10,000 bootstrapped iterations.

### Functional connectivity analysis of resting-state fMRI data

#### Condition by time interaction

For each participant, the mean time series in each seed was correlated with the time series of each gray matter voxel. Correlation coefficient *(r)* maps were examined for condition by time interactions, controlling for age and sex, using the GLM framework in CONN 20.b (SPM12, MATLAB 2018b) with 2 conditions (VRNT, Control) as ‘subject effects’, 2 timepoints as ‘conditions’) and the default canonical resting state networks as ‘seeds/sources’. The between-subject contrast for [VRNT Control condition age sex] was set to [1 -1 0 0] and the between-condition contrast for [Pre Post] was set to [-1 1]. Tests were 2-sided, with a voxel-wise cluster-forming threshold of p < 0.01 uncorr. (t > 2.68) across the whole brain, family-wise-error (FWE)-corrected for multiple comparisons at *p* < 0.05, implemented in CONN 20.b Based on prior literature linking chronic pain (and effects of treatment) to altered resting-state functional connectivity, the functional connectivity analyses were restricted to the somatomotor network (CONN toolbox seeds ‘Superior’, ‘Lateral Left’, ‘Lateral Right’), default mode network (DMN; CONN seeds ‘DMN.mpfc’, ‘DMN.pcc’), cingulo-opercular network (CONN seeds ‘aMCC’, ‘aINS Left’, ‘aINS Right’), and NAc (CONN seeds ‘Accumbens.L’, ‘Accumbens.R’).

#### Regression with pain reduction

The relationship between pre-to-post resting state connectivity and pre-to-post pain reduction was examined in the VRNT condition in networks showing a significant condition by time interaction and controlling for age and sex. The between-subject contrast for [VRNT delta_pain_VRNT age sex] was set to [0 1 0 0] and the between-condition contrast for [Pre Post] was set to [-1 1]. As above, tests were 2-sided, with a voxel-wise cluster-forming threshold of p < 0.01 across the whole brain, FWE-corrected for multiple comparisons at *p* < 0.05.

### FA analysis of white matter DWI data

#### Condition by time

Statistical analysis of the white matter FA was carried out using TBSS (Tract-Based Spatial Statistics) [72] in FSL. First, pre-post FA difference maps were created for each participant by subtracting the Pre from Post maps. TBSS was performed using *randomise*, a permutation-based inference tool for nonparametric statistical thresholding, with the number of permutations set at 5000. For condition comparisons of pre-post FA difference maps, we used GLMs controlling for age and sex, with cluster correction for multiple comparisons across the whole brain set at *p* < 0.05 (2-sided) using TFCE (threshold-free cluster enhancement) with default parameters (H = 2, E = 0.5).

#### Regression with pain reduction

The relationship between pre-post FA and pre-post pain reduction was examined in the VRNT condition in the mask of regions showing a significant condition by time effect, controlling for age and sex, and cluster-corrected across the mask at *p* < 0.05 (2-sided) using TFCE, as above.

## Results

As shown in Figure 1, we randomized 61 participants (31 male, 30 female; age *M* = 34.3 years, *SD* = 9.6; pain duration *M* = 8.5 years, *SD* = 7.1) into VRNT vs. Control, which had comparable sociodemographic characteristics across the two conditions (Table 1). Of the 31 participants randomized to VRNT, 30 completed treatment and all on-line assessments (one participant dropped out mid-treatment due to time constraints), and all 30 Control participants completed all on-line assessments. Both MRI sessions (pre- and post-) were completed by 28 VRNT participants and 24 Control participants (Fig. 1).

**Table 1.** Sample characteristics at study entry.

|  | **VRNT**<br>(*n* = 31) | **Control**<br>(*n* = 30) | **P value** |
| --- | --- | --- | --- |
| Age (yrs), *mean (SD)* | 34.8 (9.9) | 33.5 (9.2) | 0.60 |
| Sex (M, F) | 16 (53%), 15 (47%) | 15 (50%), 15 (50%) | 0.99 |
| Pain duration (yrs), *mean (SD)* | 8.7 (6.1) | 8.4 (8.1) | 0.90 |
| Race |  |  |  |
| *White* | 28 | 27 | 0.97 |
| *Black* | 1 | 2 |  |
| *Native Hawaiian or Pacific Islander* | 0 | 1 |  |
| *American Indian or Alaska Native* | 2 | 0 |  |
| *Asian* | 0 | 0 |  |
| BMI | 27.1 (5.1) | 27.0 (5.3) | 0.94 |
| Education |  |  |  |
| *High school and some college* | 15 | 15 | 0.68 |
| *College* | 8 | 10 |  |
| *More than college* | 8 | 5 |  |
| Employment status (full-time, other) |  |  |  |
| *Full-time* | 21 | 20 | 0.93 |
| *Other* | 10 | 10 |  |
| Annual income (\$ K per year), *mean (SD)* | 84.1 (94.8) | 71.6 (56.4) | |
| Current medication (past 4 weeks) <sup>a</sup> |  |  |  |
| *Over-the-counter pain relievers* | 24 | 16 | 0.71 |
| *Anxiety medication* | 1 | 3 |  |
| *Antidepressants* | 1 | 1 |  |
| *Anticonvulsants* | 2 | 1 |  |
| *Cannabinoids* | 13 | 8 |  |
| Current therapies (past 4 weeks) <sup>a</sup> |  |  |  |
| *Therapeutic exercise* | 13 | 14 | 0.97 |
| *Hands-on treatment* | 12 | 10 |  |
| *Modality treatment* | 14 | 15 |  |
| *Behavioral / mental health treatment* | 0 | 1 |  |

P-values are derived from a two-sample t-test or from a Chi-square test (across sub-categories).
<sup>a</sup> Response missing in some participants.

### Treatment effects on primary and secondary outcomes

The VRNT condition reported significantly reduced pain intensity at post-treatment vs. pre-treatment, compared to the Control condition (condition by time interaction controlled for age and sex, *g* = 0.63, medium to large effect size, *p* = 0.014). The VRNT condition averaged 35.9% ± (SD) 40.4 reduction in pain intensity vs. 11.9% ± 38.6 in the Control condition, Fig. 2B, Table 2). Clinically meaningful pain intensity reduction (i.e., at least 30% from pre-treatment) was observed in 60% (18/30) of VRNT participants compared to 30% (9/30) of Controls (*X*^2^ = 5.45, *p* = 0.019), and substantial reduction (at least 50%) was observed in 47% (14/30) of VRNT participants compared to 13% (4/30) of Controls (*X*^2^ = 7.94, *p* = 0.005).

**Table 2.**
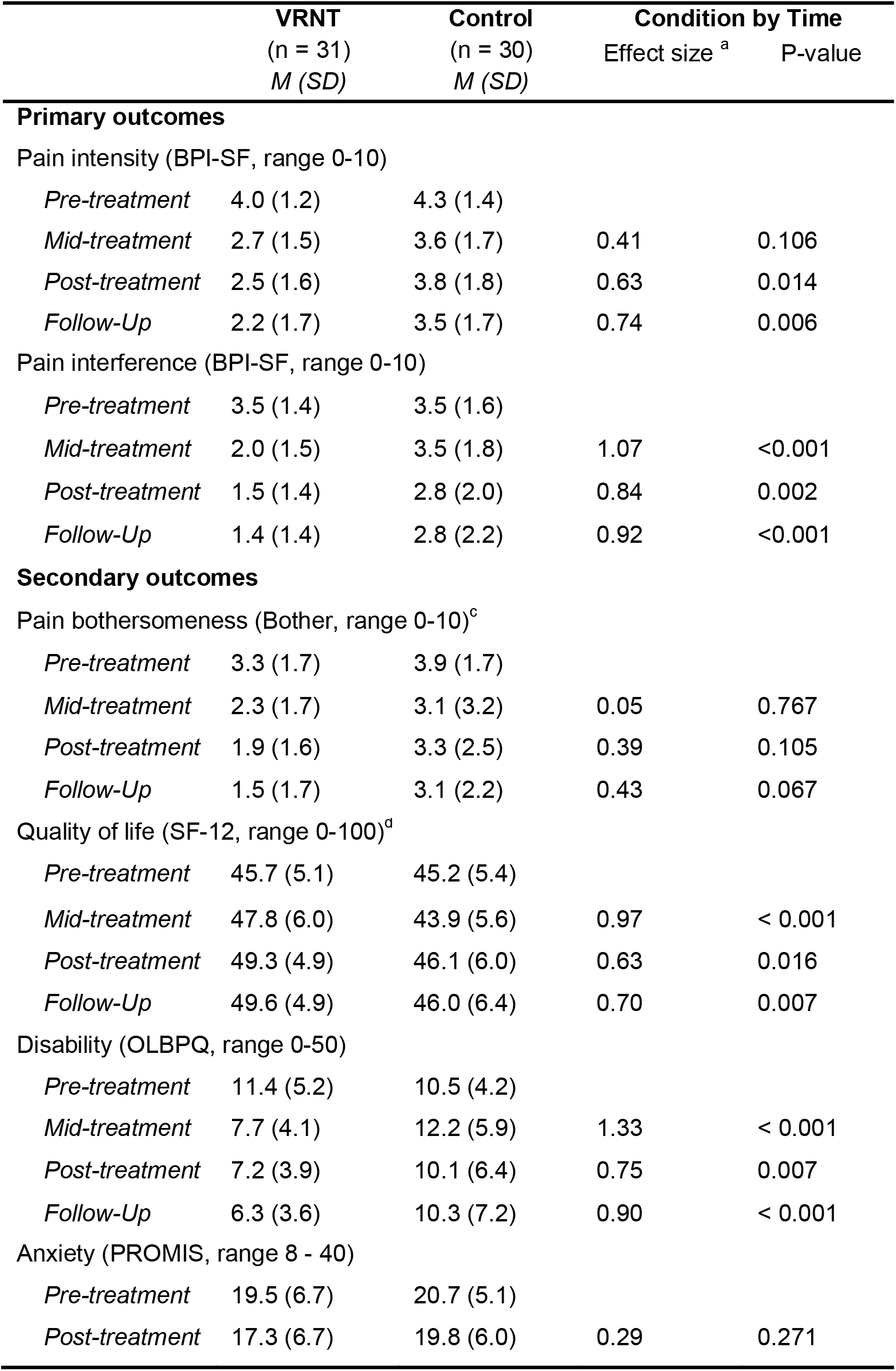
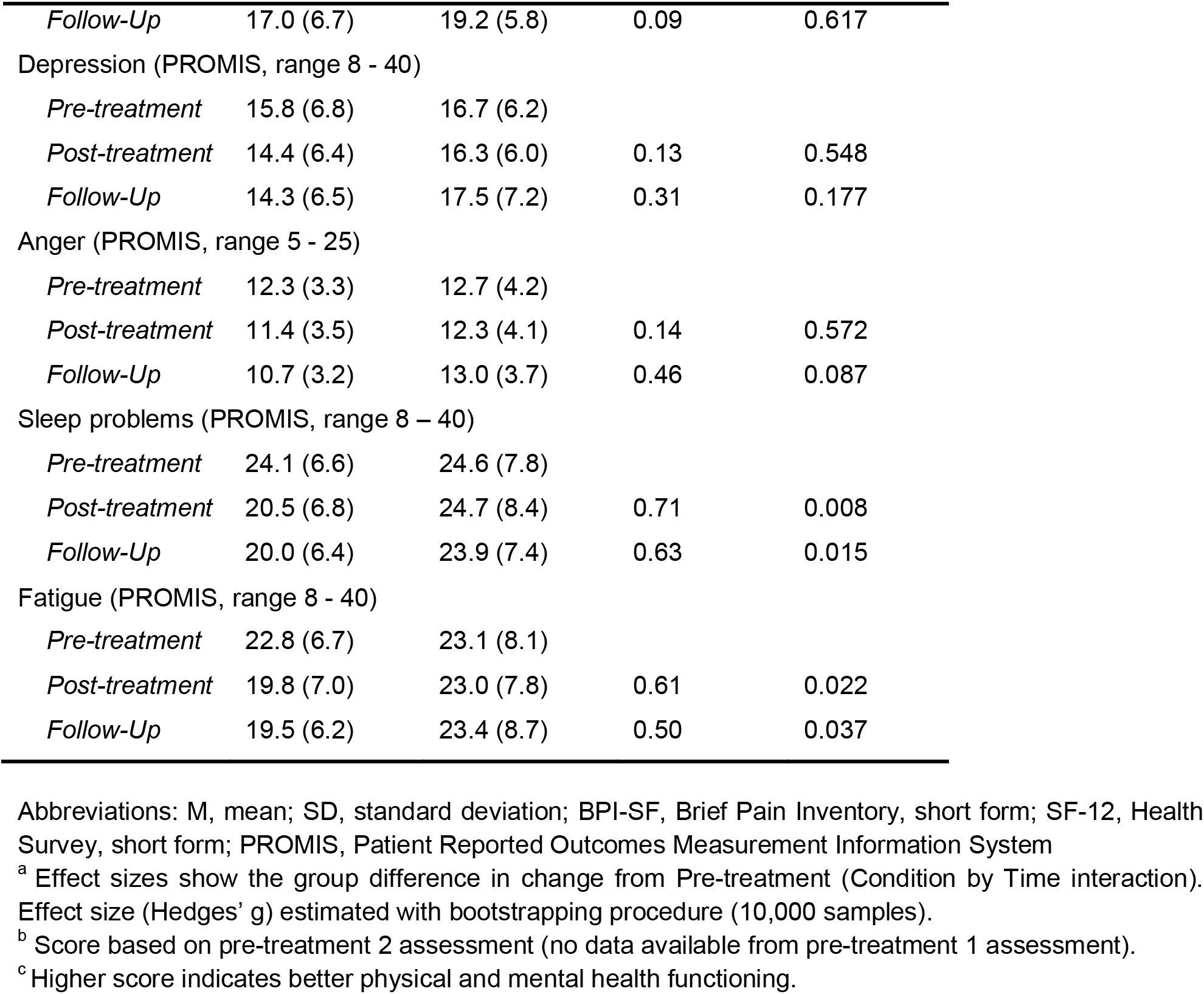
Primary and secondary clinical outcomes.

**Table 3.**
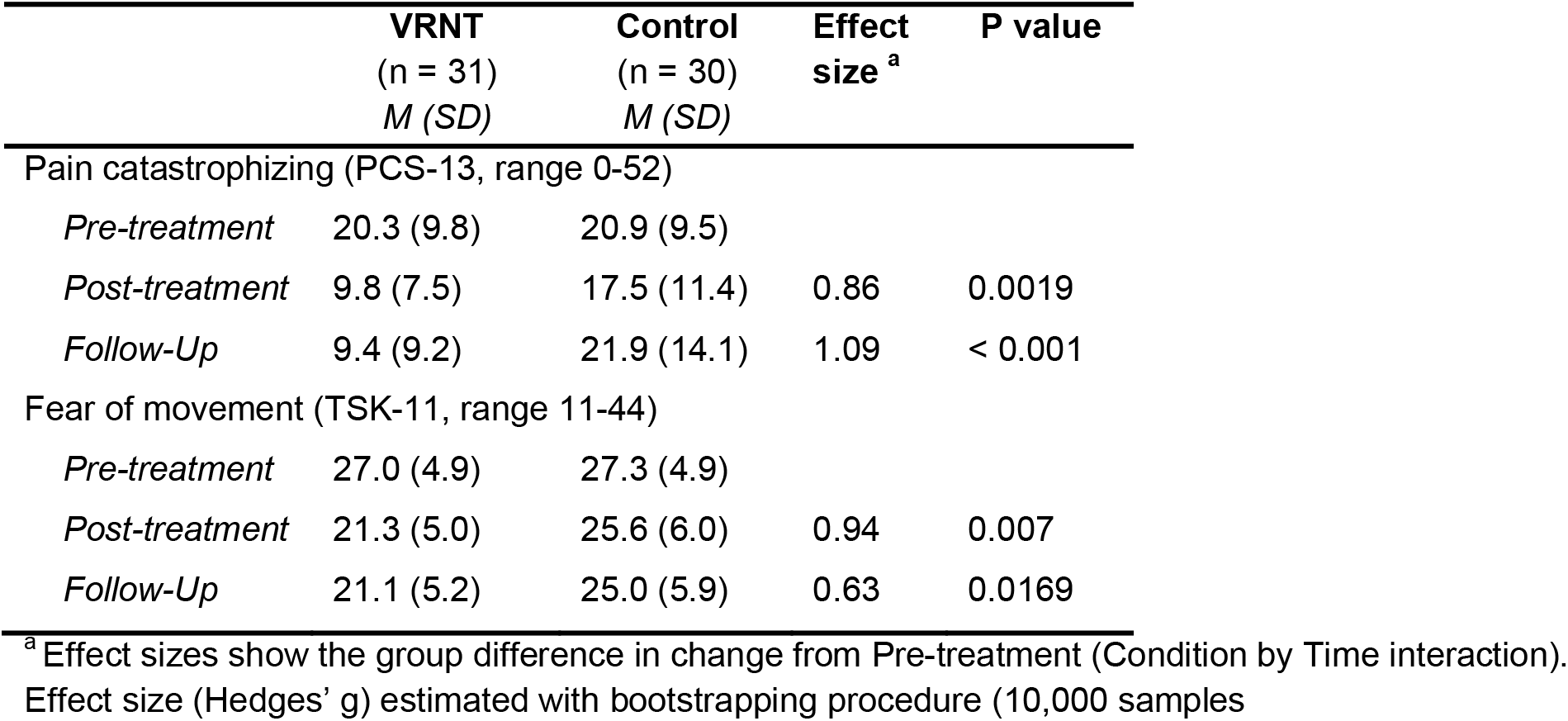
Potential mediators of treatment.

Similar to the findings with pain intensity, the VRNT condition had significantly greater reduction in pain interference at post-treatment vs. pre-treatment, compared to the Control condition (condition by time interaction controlled for age and sex *g* = 0.84, large effect size, *p* = 0.002). The VRNT condition averaged 56.3% ± 37.1 reduction in pain interference vs. 10.6% ± 73.5 reduction in the Control condition, Fig. 2C, Table 2). At least 30% reduction in pain interference was observed in 77% (23/30) of VRNT participants compared to 53% (16/30) of Controls (*X*^2^ = 3.59, *p* = 0.058), and an at least 50% reduction was observed in 60% (18/30) of VRNT participants compared to 30% (9/30) of Controls (*X*^2^ = 5.45, *p* = 0.019).

The treatment effect on both primary outcomes was already evident at mid-treatment (Table 2) and continued to improve at 2-week follow-up (slightly larger between-condition effect sizes on both primary outcomes than at post-treatment), indicating some lasting effects of VRNT and possibly continued improvement on pain outcomes (Table 2).

As shown in Table 2, the VRNT condition also had significant improvements compared to the Control condition on several secondary outcomes at post-treatment, with medium to large effect sizes (Table 2): reduced disability (condition x time interaction: *g* = 0.75, *p* = 0.01), improved quality of life (*g* = 0.63, *p* = 0.016), reduced sleep problems *(g* = 0.71, *p* = 0.008), and reduced fatigue (*g =* 0.61, *p* = 0.022). As observed above for primary outcomes, treatment effects were largely already evident at mid-treatment (if assessed) and persisted at follow-up. Pain bothersomeness showed a similar but weaker trend that was not significant (*g* = 0.39, *p* = 0.105), and the treatment effects on measures of psychological distress were not significant for anxiety (*p* = 0.271), depressive symptoms (*p* = 0.548), or anger (*p* = 0.572).

### Potential mediators of treatment

Compared to the Control condition, the VRNT condition reported significant improvements at post-treatment in pain catastrophizing (condition by time interaction controlled for age and sex: *g* = 0.86, *p* = 0.002) and kinesiophobia (*g* = 0.94, *p* = 0.007), with large effects. Though kinesiophobia and pain catastrophizing were significantly correlated (at baseline *r* = 0.56, *p* < 0.001; pre- to post-treatment change scores *r* = 0.52, *p* < 0.001), suggesting they are partially overlapping constructs, their change scores were differentially related to changes on the primary outcomes (pain intensity, pain interference), regardless of condition. Across the full sample (VRNT and Control conditions combined), the change in kinesiophobia correlated positively with the change in pain intensity (*r* = 0.42, *p* = 0.001) and in pain interference (*r* = 0.54, *p* < 0.001), whereas the change in pain catastrophizing correlated positively with the change in pain interference only (*r* = 0.35, *p* = 0.008; correlation with pain intensity *r* = 0.15, *p* = 0.280). For the significant correlations with primary outcomes in the full sample, we next tested whether changes in kinesophobia and pain catastrophizing mediated the effect of VRNT vs. Control on changes in the relevant primary outcomes. The mediation path VRNT vs. Control → Δ TSK → Δ pain intensity was significant; that is, the reduction in kinesiophobia fully mediated the reduction in pain intensity from VRNT (β_ab_ = -0.24, *p* = 0.031; β_c_ (direct effect) = -0.53, *p* = 0.009, β_c’_ (controlling for mediator) = -0.29, *p* = 0.178 Fig. 2E). The reduction in kinesiophobia also fully mediated the reduction in pain interference from VRNT (VRNT vs. Control → Δ TSK → Δ pain interference: β_ab_ = -0.35, *p* = 0.002; β_c_ (direct effect) = -0.69, *p* = 0.001, β_c’_ (controlling for mediator) = -0.35, *p* = 0.127). Finally, the reduction in pain catastrophizing partially mediated the reduction in pain interference from VRNT (VRNT vs. Control → Δ PCS → Δ pain interference: β_ab_ = -0.16, *p* = 0.048; β_c_ (direct effect) = -0.69, *p* < 0.001, β_c’_ (controlling for mediator) = -0.53, *p* = 0.029).

### Treatment effects on brain outcomes

Of participants who had available pre- and post-MRI data, all were included in the resting-state analysis, and one participant’s data was excluded from the white-matter analysis, for final included samples of: resting-state analysis, 28 VRNT, 24 Control; white-matter analysis, 28 VRNT, 23 Control.

### Resting-state fMRI functional connectivity

We tested treatment effects on functional connectivity of brain networks linked to chronic back pain and treatment including the somatomotor network (superior and lateral aspects), DMN (PCC and MPFC), and cingulo-opercular network (aMCC and insula). We observed significant condition by time interaction (controlled for age and sex) in functional connectivity of the somatomotor network and DMN, with the VRNT condition showing in both networks increased resting-state connectivity after treatment vs. control condition (Table 4). Specifically, in the VRNT condition the somatomotor network had significantly increased connectivity with prefrontal cortical areas (Fig. 3A-C), including bilateral dorsal and medial prefrontal cortices (dLPFC, dMPFC), anterior cingulate (aMCC), bilateral inferior parietal lobule (IPL), bilateral middle temporal gyrus (MTG), and cerebellum (all p < 0.05 FWE-corr., see Table 4 for exact values). The posterior DMN (PCC) had significantly increased connectivity after treatment vs. control condition with the dLPFC (p < 0.05 FWE-corr.; Fig. 3D, Table 4 and the anterior DMN (MPFC) with the anterior PFC / frontal pole (approaching significance at p = 0.055 FWE-corr., Table 4). No significant condition by time interaction effects (controlled for age and sex) were observed for the connectivity of the cingulo-opercular network. We also tested the connectivity of the NAc, to examine any treatment effects on the frontostriatal (NAc-MPFC) circuitry implicated in chronic pain. No significant condition by time interactions effects (controlled for age and sex) were observed for the connectivity of either left or right NAc with MPFC. Instead, left NAc had significantly decreased connectivity after treatment vs. control with the right IPL and the left middle occipital gyrus and (both p < 0.05 FWE-corr., see Table 4).

**Figure 3.**
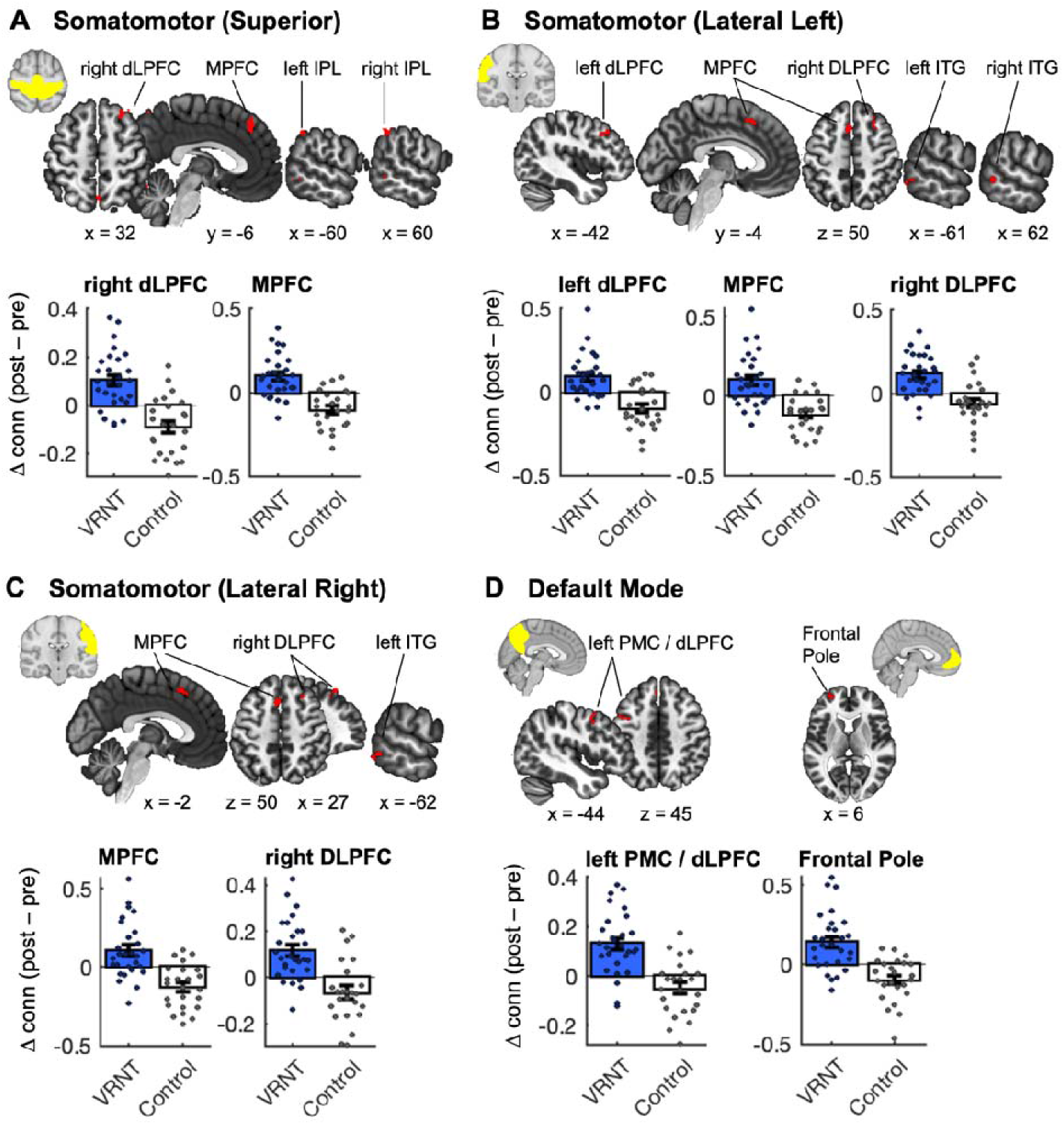
Treatment effects on functional connectivity. **(A) – (D)** Brain maps show clusters of significant condition by time interaction, controlled for age and sex, of brain-wide correlation coefficient (‘functional connectivity’) maps for each ROI; p < 0.05 FWE-corrected; 2 x 2 ANOVA with two conditions (VRNT, Control) and 2 timepoints (Pre, Post); computed with CONN toolbox (20.b; SPM 12, MATLAB 2018b). Bar plots show connectivity post-pre difference scores between ROI and selected regions; dots are individual scores; bars are mean and standard error; blue, VRNT; white, Control. dLPFC, dorsolateral prefrontal cortex; MPFC, medial prefrontal cortex; IPL, inferior parietal lobule; ITG, inferior temporal gyrus; PMC, premotor cortex; L, left; R, right.

**Table 4.**
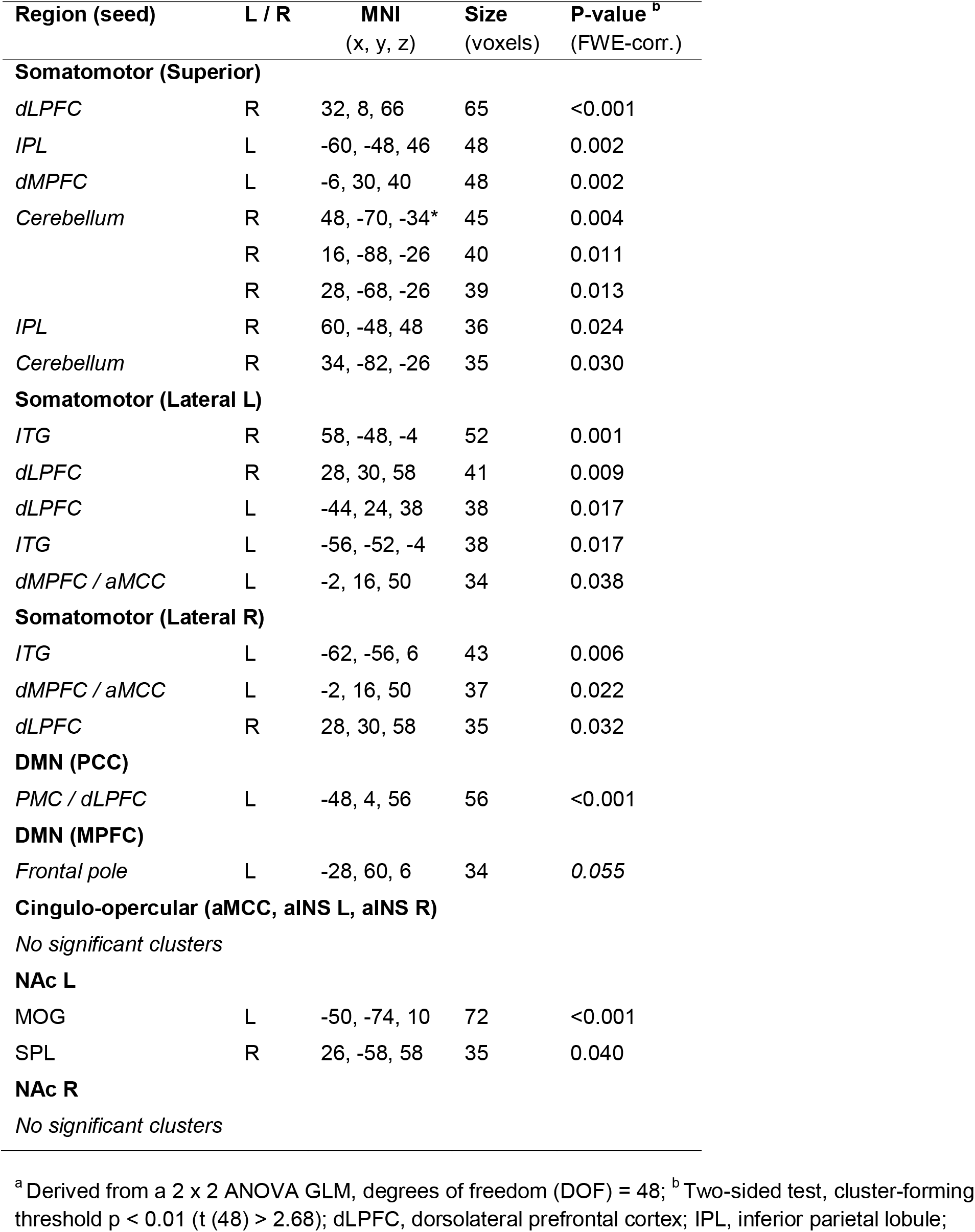
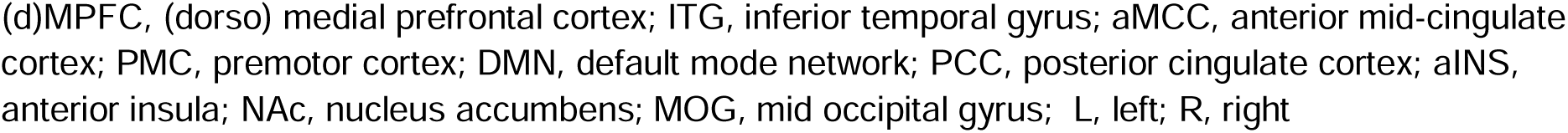
Resting-state functional connectivity, significant group by time interaction results.

| Region (seed) | L / R | MNI<br>(x, y, z) | Size<br>(voxels) | P-value<sup>b</sup><br>(FWE-corr.) |
| --- | --- | --- | --- | --- |
| **Somatomotor (Superior)** |  |  |  |  |
| *dLPFC* | R | 32, 8, 66 | 65 | <0.001 |
| *IPL* | L | -60, -48, 46 | 48 | 0.002 |
| *dMPFC* | L | -6, 30, 40 | 48 | 0.002 |
| *Cerebellum* | R | 48, -70, -34* | 45 | 0.004 |
|  | R | 16, -88, -26 | 40 | 0.011 |
|  | R | 28, -68, -26 | 39 | 0.013 |
| *IPL* | R | 60, -48, 48 | 36 | 0.024 |
| *Cerebellum* | R | 34, -82, -26 | 35 | 0.030 |
| **Somatomotor (Lateral L)** |  |  |  |  |
| *ITG* | R | 58, -48, -4 | 52 | 0.001 |
| *dLPFC* | R | 28, 30, 58 | 41 | 0.009 |
| *dLPFC* | L | -44, 24, 38 | 38 | 0.017 |
| *ITG* | L | -56, -52, -4 | 38 | 0.017 |
| *dMPFC / aMCC* | L | -2, 16, 50 | 34 | 0.038 |
| **Somatomotor (Lateral R)** |  |  |  |  |
| *ITG* | L | -62, -56, 6 | 43 | 0.006 |
| *dMPFC / aMCC* | L | -2, 16, 50 | 37 | 0.022 |
| *dLPFC* | R | 28, 30, 58 | 35 | 0.032 |
| **DMN (PCC)** |  |  |  |  |
| *PMC / dLPFC* | L | -48, 4, 56 | 56 | <0.001 |
| **DMN (MPFC)** |  |  |  |  |
| *Frontal pole* | L | -28, 60, 6 | 34 | 0.055 |
| **Cingulo-opercular (aMCC, aINS L, aINS R)** |  |  |  |  |
| *No significant clusters* |  |  |  |  |
| **NAc L** |  |  |  |  |
| *MOG* | L | -50, -74, 10 | 72 | <0.001 |
| *SPL* | R | 26, -58, 58 | 35 | 0.040 |
| **NAc R** |  |  |  |  |
| *No significant clusters* |  |  |  |  |

<sup>a</sup> Derived from a 2 x 2 ANOVA GLM, degrees of freedom (DOF) = 48; <sup>b</sup> Two-sided test, cluster-forming threshold $p < 0.01$ ( $t(48) > 2.68$ ); dLPFC, dorsolateral prefrontal cortex; IPL, inferior parietal lobule;
(d)MPFC, (dorso) medial prefrontal cortex; ITG, inferior temporal gyrus; aMCC, anterior mid-cingulate cortex; PMC, premotor cortex; DMN, default mode network; PCC, posterior cingulate cortex; aINS, anterior insula; NAc, nucleus accumbens; MOG, mid occipital gyrus; L, left; R, right

### White matter FA

We observed several clusters showing significant condition by time interaction (controlled for age and sex) in FA in the white matter adjacent to the aMCC (largely in the right corpus callosum, Table 5), with the VRNT condition showing decreased FA with treatment as compared to the Control condition (Fig. 4A). Importantly, within this region, we observed a cluster where the pre-to-post FA reduction t in the VRNT condition was significantly positively correlated with the pre-to-post pain reduction (Fig. 4B).

**Figure 4.**
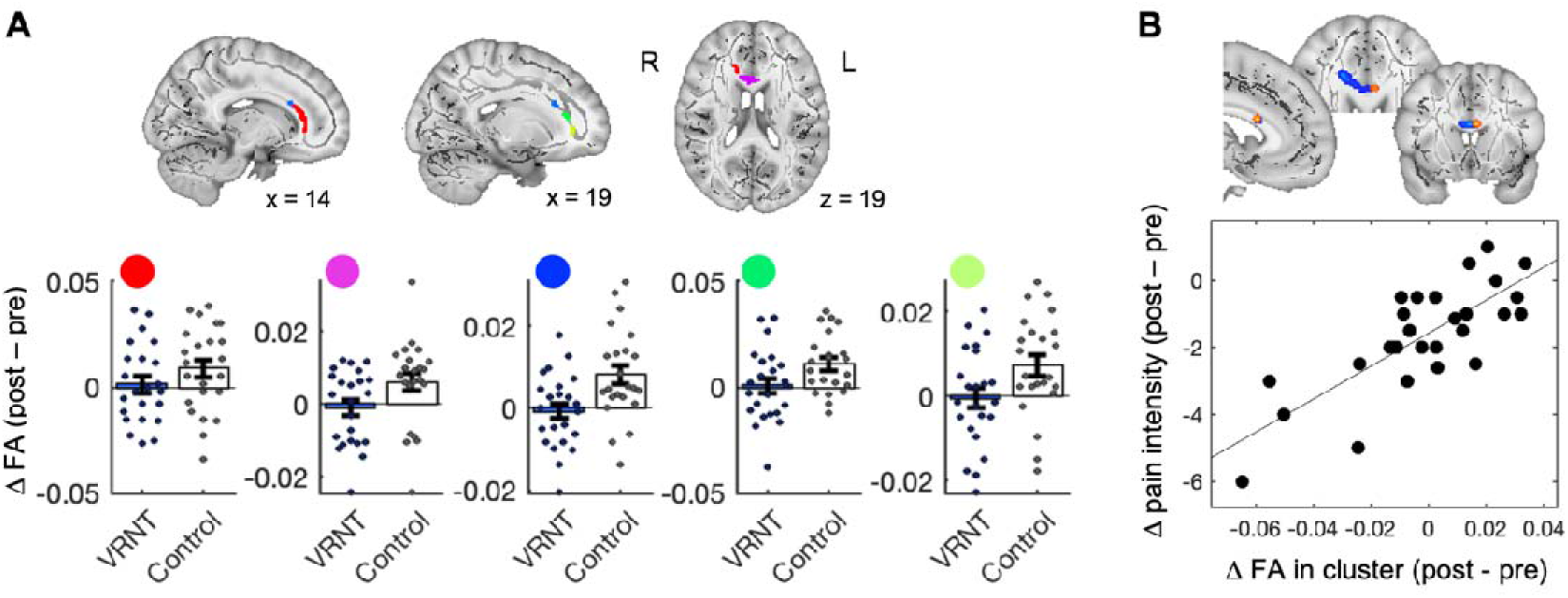
Treatment effects on white matter fractional anisotropy (FA). Brain maps show clusters of significant condition by time interaction, controlled for age and sex, of white-matter FA tested across the entire white matter skeleton; p < 0.05 TBSE-corrected; 2-sided t-test of difference maps (VRNT(Post-Pre), Control (Post-Pre)) computed with TBSS in FSL. Bar plots show FA post-pre difference scores; dots are individual scores; bars are mean and standard error; blue, VRNT; white, Control. (E) Scatter plot shows for each individual participant the relationship between the pain intensity post-pre difference score and FA post-pre difference scores in the area of significant condition by time interaction, controlled for age and sex. L, left; R, right.

**Table 5.** White matter results.

| Region | L / R | MNI<br>(x, y, z) | Size<br>(voxels) | Peak<br>t-value<sup>a</sup> | P-value<sup>b</sup><br>(TFCE-corr.) |
| --- | --- | --- | --- | --- | --- |
| **Group by time interaction** |  |  |  |  |  |
| *Corpus callosum* | R | 14, 30, 13 | 86 | -2.29 | < 0.05 |
|  | R | 4, 19, 17 | 48 | -2.73 | < 0.05 |
|  | R | 17, 25, 22 | 32 | -3.06 | < 0.05 |
|  | R | 19, 34, 14 | 14 | -2.58 | < 0.05 |
|  | R | 19, 40, 1 | 13 | -2.88 | < 0.05 |
| **Correlation with pain reduction in the VRNT group<sup>c</sup>** |  |  |  |  |  |
| *Corpus callosum* | L | -4, 16, 20 | 3 | 6.03 | < 0.05 |

<sup>a</sup> Value derived from unpaired t-test of difference maps (VRNT condition (Post-Pre) – Control condition (Post-Pre)); <sup>b</sup> Two-sided test; <sup>c</sup> examined for regions showing significant group by time interaction.

## Discussion

We tested the effects of a VR behavioral intervention (VRNT) on clinical and brain outcomes in patients with CBP. Compared to usual care/waitlisted controls, VRNT led to significantly reduced pain intensity and pain interference at post-treatment, which persisted at 2-week follow-up, and these effects were partially mediated by a reduction in kinesiophobia and pain catastrophizing. Several secondary outcomes were also improved, including disability, quality of life, sleep, and fatigue. In addition, VRNT was associated with significant brain changes compared to the control condition.

VRNT demonstrated clinically substantial reductions in pain intensity and pain interference, with medium to large effect sizes. Nearly half of participants receiving VRNT obtained 50% or greater pain reduction (compared to only 13% of controls), and VRNT doubled the frequency of obtaining 50% or greater reduction in pain interference (60% vs. 30% in controls). These effects appear to exceed the effects of leading behavioral / psychological therapies - CBT and acceptance and mindfulness-based therapies [28,31,60,85,92]). The limited efficacy of these approaches may be due to the fact they are not guided by contemporary conceptual models of pain that place the brain as the centerpiece of a changeable - even potentially reversible - chronic pain experience, and their exercises / techniques may not be experientially powerful. Newer therapies based on this model - such as PRT and EAET - have yielded substantial pain reduction [4,51], including superior outcomes to CBT on some pain-related variables in two trials [52,94]. This suggests that interventions targeting the brain’s role in chronic pain, as in VRNT, by changing one’s attributions for the etiology of the pain (from body to brain) and reducing pain-related and broader emotional and interpersonal fears, may lead to greater pain reduction compared to conventional behavioral therapies. There also is hope that VR interventions for chronic pain will yield better longer-term outcomes than the traditional approaches [21,30,77,86], which often exhibit small follow-up effects [92]. Although our test of VRNT had only a 2-week follow-up - and future studies warrant a longer follow-up - our preliminary results suggest maintained or continued gains in pain-related outcomes.

Even newer psychological therapies face limitations due to their reliance on the availability, training, and skills of therapists, as well as patient access and comfort with interpersonal, face-to-face treatments. Both newer and traditional therapies are also limited in the types of experiential exercises they can provide. VRNT appears to overcome these limitations, by combining pain psychology and neuroscience with unique first-person experiential learning. The pain neuroscience component educates patients about the brain-based etiology of their pain and its potential for substantial change through actions, cognitions, and emotions. VRNT’s active learning experiences with a personalized avatar and audiovisual representation of pain help patients shift from fearful attributions of bodily causes of their pain to changeable central causes, Moreover, VRNT is patient-led and can be used at home, making it more accessible and overcoming feasibility barriers of in-person, therapist-led treatments. Notably, our study was conducted during the COVID pandemic, testing VRNT’s accessibility in a real-world scenario, with all interactions with study personnel either conducted virtually (educational video, training session, weekly calls) or heavily restricted and regulated (MRI scanning under strict COVID protocols). Despite these restrictions, participants were able to follow the study procedures, and participant retention was nearly perfect.

VRNT showed significant improvements in pain and functioning but did not lead to significant changes in psychological distress (depressive symptoms, anxiety, anger), at least at 2 weeks after treatment. Similar outcomes has been observed with EAET [51] and a pain neuroscience-related intervention [41], suggesting that achieving improvements in negative affect might be more challenging, delayed, or less reliable with these newer interventions. The strong focus on changing attributions and fears about pain’s etiology, without specific techniques or components to improve mood, could explain improved pain-related outcomes but lack of effect on negative affect. To achieve improved affect, longer VRNT interventions, more specialized intervention components targeting affect specifically, or longer follow-up see if the improved pain is followed by improved affect may be needed.

To investigate purported neural mechanisms of action, we conducted pre and post MRI, hypothesizing that VRNT would affect structural and functional connectivity in brain networks most consistently associated with chronic pain and treatment effects, including the somatomotor network, default mode network (DMN), cingulo-opercular network, and frontostriatal circuitry. VRNT (vs. control) was associated with modest increases (at a fairly liberal statistical threshold) in functional connectivity of the somatomotor network and DMN after treatment, largely with prefrontal cortical areas (dLPFC, dMPFC / aMCC and aPFC / frontal pole). No significant effects were observed for the cingulo-opercular and frontostriatal circuitry at that threshold.

These findings align with previous neuroimaging research on successful behavioral interventions for pain, indicating increased prefrontal-somatosensory functional connectivity [4] and gray matter volume [68], and increased pain-related recruitment of prefrontal cortices [33] Prefrontal and somatomotor cortices are commonly implicated in musculoskeletal chronic pain [9,67], with dLPFC, mPFC, and primary somatomotor cortex being among the regions showing the most robust changes with prolonged pain and the most promising effects of non-pharmacological pain interventions [4,11,32,33,37,40,44,45,54,64,68,69,78,91], suggesting that these treatments might result in neurobiologically detectable improvements. In addition, some recent studies using non-invasive brain stimulation have successfully targeted left dLPFC and primary somatomotor cortices to alleviate musculoskeletal pain [1,66], cf. [3], lending further support to the key roles these areas play in chronic pain and recovery [34]. Together with earlier work, our findings may reflect increased top-down regulatory pain control via the dLPFC (paralleling the observed reductions in pain intensity and interference with VRNT vs. control) and altered (‘normalized’) somatosensory processing of noxious (and non-noxious) input - away from the threatening words like ‘pain’ towards more benign terms like ‘sensation.’ Such a shift in language to describe the patients’ experience was noted anecdotally in some VRNT participants in the current study, and was found in qualitative analyses of the reports of back pain patients receiving PRT [79].

We did not observe any treatment effects in the MPFC-NAc connectivity, previously implicated in the transition to chronic pain [7,26] and cognitive regulation of evoked pain [93]. Recent studies have reported mixed findings and lack of treatment effects in this particular neural circuitry [4,58,59], suggesting it might not be a reliable marker of chronic pain. Instead, accumulating evidence points to a more comprehensive cortico-limbic circuitry involvement in chronic pain, including the findings of increased VRNT-related connectivity of superior parietal and visual cortices with the NAc in the present study.

Studies of white matter changes in musculoskeletal chronic pain have produced inconclusive results [25], reporting decreases [10,36,48,88], increases or no change [11,20,46,53,76] of white matter FA values in pain patients. Though inconsistent, these findings are often localized to the corpus callosum, anterior cingulum, and frontal white matter, which aligns with our significant differences in VRNT vs. control. Our findings were driven by an increase in FA post vs. pre-treatment in the control condition, which was not observed in the VRNT condition, making the condition difference difficult to interpret. However, the lack of FA increase in the VRNT condition was associated with pain reduction. VRNT participants experiencing the greatest pain relief had the most unchanged FA values post relative to pre-treatment, suggesting that the preserved FA might be clinically relevant. Interventions for chronic pain might potentially affect white matter and normalize maladaptive neuroplastic changes associated with persistent pain, as also indicated by preliminary results in chronic pain patients post-surgery [11,46]. Nevertheless, the neurobiological relevance of white matter findings and treatment effects in musculoskeletal pain remains unclear.

This study has several limitations. The sample size, although adequately powered to identify the medium-large effects that were observed, is still relatively small. Although the sample was balanced in gender, it was restricted in race to almost all White participants. Replication with a larger, more racially/ethnically diverse sample, and with a longer follow-up time is needed. This trial had a passive control, and replication with an active, placebo control, such as has been done with another successful VR approach for chronic pain [22] is needed to further specify VRNT’s effects. Finally, our functional connectivity analyses were performed at a liberal corrected statistical threshold and restricted to hypothesis-driven regions and networks of interest (as in most other studies) and therefore likely biased by prior positive research findings.

In conclusion, the novel VR-based treatment for chronic musculoskeletal pain, VRNT, showed preliminary efficacy in significantly improving pain-related outcomes, and it may do so via changes in somatosensory and prefrontal brain networks. Our results contribute to the literature on the value of VR for chronic pain and offer an approach based on newer principles of brain-based, reversible pain and novel VR experiential components – a personalized avatar and changeable pain representations. Dissemination and implementation of VRNT thus has the potential for substantial impact on the epidemic of chronic pain.

## Data Availability

All data produced in the present study are available upon reasonable request to the authors

## Acknowledgements

This research was supported by the National Institutes of Health under the NIH HEAL Initiative under award number R43NR017575 to T.A.B., M.C., and L.W.

