## Supplemental Material for "The Effects of Virtual Reality Neuroscience-based Therapy on Clinical and Neuroimaging Outcomes in Patients with Chronic Back Pain: A Randomized Clinical Trial"

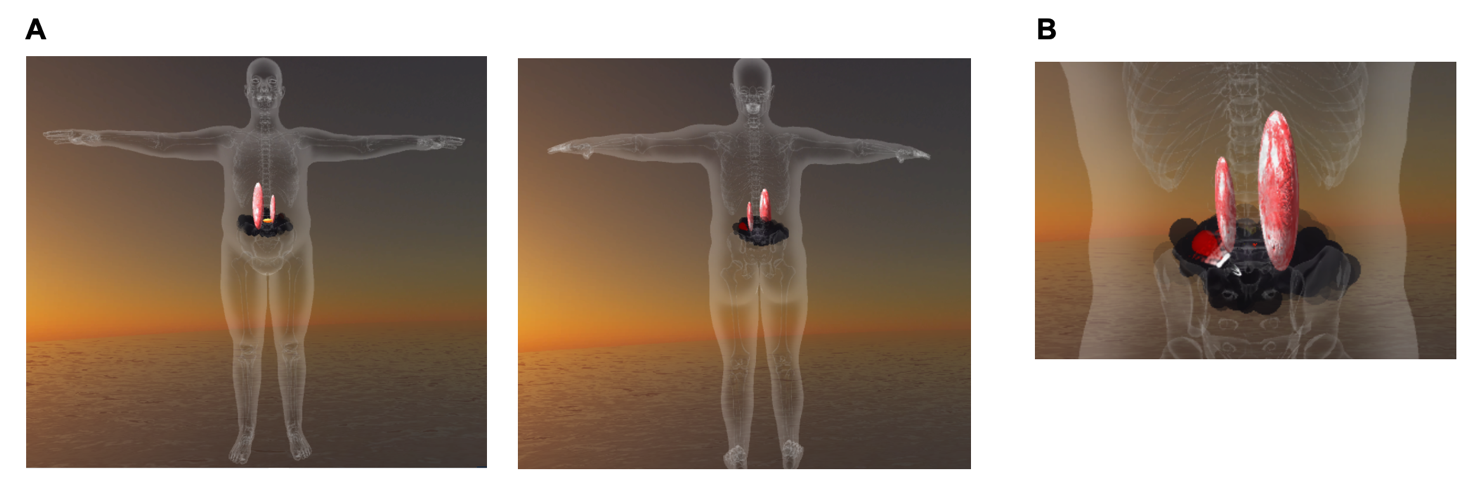

**Supplementary Figure 1.** An example of a finalized custom pain avatar in the mobile VR environment, created by the study participant with assistance by the study team member. A) front and back view of the avatar (selected from 9 options varying in gender, body shape, body size). **B)** individualized representation of pain experience, personalized in terms of shape, color, movement / animation and sound.

**Supplementary Methods**

**Other potential mediators**

Other potential mediators of treatments assessed in this study included attitudes towards pain, self-efficacy, optimism, meaning and purpose in life, mindfulness, and emotion regulation capacity.

Attitudes towards pain were assessed using the Fear of Pain Questionnaire (FOP, [10]) and the Short-Form Survey of Pain Attitudes, Emotion subscale (SOPA-Emo, [15]). The sense of self-efficacy was assessed using the Generalized Self-Efficacy Scale (GSE, [14]). Dispositional optimism was assessed using the Revised Life Orientation Test (LOT-R, [6]. Meaning and purpose in life were assessed using the PROMIS Meaning and Purpose measure (form 4a, [12]). Mindfulness was assessed using the Mindfulness and Attention Awareness Scale (MAAS, [4]). Emotion regulation capacity was assessed using the Emotion Regulation Questionnaire (ERQ, [8])

**MRI preprocessing pipelines**

Anatomical data. T1-weighted (T1w) images were corrected for intensity non-uniformity (INU) using *N4BiasFieldCorrection* [17] and used as T1w-reference throughout the workflow. The T1w-reference was then skull-stripped using ANTs 2.2.3 with OASIS30ANTs as target template. Brain tissue segmentation of cerebrospinal fluid (CSF), white matter (WM) and gray matter (GM) was performed on the brain-extracted T1w using *fast* (FSL 6.0.5, [19]). Volume-based spatial normalization to the *ICBM 152 Nonlinear Asymmetrical template version 2009c* [5] was performed through nonlinear registration with antsRegistration (ANTs 2.3.3), using brain-extracted versions of both T1w reference and the T1w template.

Resting-state fMRI data. A *B0*-nonuniformity map (*fieldmap*) was estimated using *topup* (FSL 6.0.5, ([2]). A reference volume and its skull-stripped version were generated by aligning and averaging 1 single-band reference image (SBRefs). Head motion parameters (transformation matrices, and six corresponding rotation and translation parameters) were estimated with respect to the BOLD reference before any spatiotemporal filtering using *mcflirt* (FSL 6.0.5, [9]). The estimated *fieldmap* was then aligned using rigid-body registration to the target BOLD reference run. The field coefficients were mapped onto the BOLD reference using the transform. The BOLD reference was then co-registered to the T1w reference using *bbregister* with 6 degrees of freedom (FreeSurfer; boundary-based registration [7]). The confounds included time series derived from head motion estimates (framewise displacement (FD) and DVARS [9,11] and global signals (CSF, WM, whole-brain), as well as temporal derivatives and quadratic terms for each [13]). Frames that exceeded a threshold of 0.5 mm FD or 1.5 standardized DVARS were annotated as motion outliers (spikes). The BOLD time-series were resampled into standard space using *antsApplyTransforms* (ANTs).

White matter DWI data. A total of 4 DWI series were concatenated, with the following preprocessing operations performed on individual DWI series before concatenation. Any images with a b-value less than 100 s/mm^2^ were treated as a b=0 image. MP-PCA denoising as implemented in MRtrix3's *dwidenoise* [16,18] was applied with a 5-voxel window and field inhomogeneity corrected using *dwibiascorrect* from MRtrix3 with the N4 algorithm. Next, the mean intensity of the DWI series was adjusted across the 4 DWI scanning sequences. FSL (6.0.3)'s *eddy* was used for head motion correction and Eddy current correction [3] with $q$-space smoothing factor of 10, 5 iterations, and 1000 voxels to estimate hyperparameters. Linear first and second level models were used to characterize Eddy current-related spatial distortion. $q$-space coordinates were assigned to shells. Field offset was attempted to be separated from subject movement. Shells were aligned post-eddy. Eddy's outlier replacement was run [1]. Data were grouped by slice, only including values from slices with at least 250 intracerebral voxels. Groups deviating by more than 4 SD from the prediction had their data replaced with imputed values. Data was collected with reversed phase-encode blips, resulting in pairs of images with distortions going in opposite directions. Here, b=0 reference images with reversed phase encoding directions were used along with an equal number of b=0 images extracted from the DWI scans. From these pairs the susceptibility-induced off-resonance field was estimated [2]. The fieldmaps were ultimately incorporated into the Eddy current and head motion correction interpolation. Final interpolation was performed using the `jac` method. Framewise displacement was calculated as above for functional data [9,11], and slicewise cross correlation was also calculated. The DWI time-series were resampled to ACPC with 2mm isotropic voxels.

**Supplementary Table 1**. Other potential mediators of treatment.

|  | **VRNT**  n = 31  *mean (SD)* | **Control**  n = 30  *mean (SD)* | **Effect size ^a^** | **P value ^b^** | **% Improved from**  **Pre-treatment**  *mean (SD)* | |
| --- | --- | --- | --- | --- | --- | --- |
|  |  |  |  |  | **VRNT** | **Control** |
| Fear of pain (FOP) | | | | |  |  |
| *Pre-treatment* | 81.65 (15.22) | 77.27 (17.67) |  |  |  |  |
| *Post-treatment* | 75.31 (15.91) | 76.86 (15.85) | 0.60 | 0.031 | 8.4 (15.8) | -1.2 (14.1) |
| *Follow-Up* | 75.67 (18.46) | 72.86 (19.13) | 0.19 | 0.489 |  |  |
| Pain attitudes (SOPA-Emo) | | | | |  |  |
| *Pre-treatment* | 5.05 (1.78) | 5.28 (1.98) |  |  |  |  |
| *Post-treatment* | 5.45 (2.43) | 5.24 (1.99) | 0.16 | 0.483 | -12.14 (55.37) | -11.3 (67.4) |
| *Follow-Up* | 5.63 (2.44) | 5.00 (2.33) | 0.43 | 0.098 |  |  |
| Self-efficacy (GSE) | | | | |  |  |
| *Pre-treatment* | 31.94 (4.08) | 32.52 (3.65) |  |  |  |  |
| *Post-treatment* | 33.14 (4.36) | 32.28 (4.72) | 0.34 | 0.171 | -4.2 (9.7) | -0.3 (13.4) |
| *Follow-Up* | 33.30 (4.09) | 31.48 (4.11) | 0.78 | 0.003 |  |  |
| Dispositional optimism (LOT-R) ^c^ | | | | |  |  |
| *Pre-treatment* | 15.04 (4.14) | 13.40 (4.72) |  |  |  |  |
| *Post-treatment* | 15.00 (3.98) | 13.75 (4.44) | 0.25 | 0.385 | -1.2 (16.9) | -5.9 (19.7) |
| *Follow-Up* | 15.15 (4.98) | 12.35 (4.96) | 0.06 | 0.699 |  |  |
| Meaning and purpose in life (PROMIS) ^d^ | | | | | | |
| *Pre-treatment* | 14.43 (2.99) | 13.81 (3.56) |  |  |  |  |
| *Post-treatment* | 15.62 (3.10) | 13.86 (3.79) | 0.38 | 0.203 | -9.4 (20.5) | -3.4 (34.8) |
| Mindfulness (MAAS) ^c^ | | | | |  |  |
| *Pre-treatment* | 3.55 (0.78) | 3.53 (0.94) |  |  |  |  |
| *Post-treatment* | 3.52 (0.75) | 3.55 (0.93) | 0.02 | 0.979 | 1.2 (16.3) | 1.8 (16.8) |
| *Follow-Up* | 3.63 (0.83) | 3.54 (1.02) | 0.39 | 0.356 |  |  |
| Emotion regulation capacity – regulation (ERQ-R) ^c^ | | | | | | |
| *Pre-treatment* | 4.49 (1.07) | 4.63 (1.11) |  |  |  |  |
| *Post-treatment* | 5.21 (0.98) | 4.67 (1.30) | 0.74 | 0.009 | -17.3 (23.2) | -1.8 (18.7) |
| *Follow-Up* | 5.03 (1.12) | 4.41 (0.88) | 0.62 | 0.023 |  |  |
| Emotion regulation capacity – suppression (ERQ-S) ^c^ | | | | | | |
| *Pre-treatment* | 3.49 (1.33) | 3.81 (1.26) |  |  |  |  |
| *Post-treatment* | 3.77 (1.15) | 3.60 (1.34) | 0.24 | 0.239 | -21.5 (52.6) | -6.5 (41.7) |
| *Follow-Up* | 3.89 (1.25) | 3.79 (1.14) | 0.46 | 0.136 |  |  |

Abbreviations: FOP, Fear of Pain; SOPA-Emo, Survey of Pain Attitudes – Emotion subscale; GSE, General Self-Efficacy; LOT-R, Life Orientation Test Revised; PROMIS, Patient Reported Outcomes Measurement Information System; MAAS, Mindful Attention Awareness Scale; ERQ, Emotion Regulation Questionnaire.

^a^ Effect sizes show the group difference in change from Pre-treatment (Group by Time interaction). Effect size (Hedges’ g) was estimated using a bootstrapping procedure (n = 10,000).

^b^ p-values associated with the Group by Time interaction.

^c^ Score based on pre-treatment 2 assessment (no pre-treatment 1 assessment).

^d^ No pre-treatment 2 assessment; no follow-up.
